# Long-term Serial Exercise Stress Testing in Catecholaminergic Polymorphic Ventricular Tachycardia on Beta-Blocker and Flecainide Therapy

**DOI:** 10.1101/2025.04.08.25325493

**Authors:** Fernando Wangüemert-Perez, German Ostos-Cañero, Marta Wangüemert-Guerra, Carlos Acosta-Materan, Aridane Cárdenes-león, Eduardo Caballero-Dorta, Kevin Pérez-Rodríguez, Ramón Brugada, Josep Brugada, Efrén Martínez-Quintana

## Abstract

**Introduction:** Catecholaminergic Polymorphic Ventricular Tachycardia (CPVT) is a potentially life-threatening arrhythmic disorder typically treated with beta-blockers and, occasionally, with flecainide.

**Methods:** All patients underwent genetic testing, ECG, echocardiogram, and exercise tests. Ventricular arrhythmias were assessed using qualitative and quantitative scoring. Flecainide dosing was gradually increased, and follow-up extended from 2007 to 2024.

**Results:** 235 patients were genetically positive for the *RyR2 p*.*Gly357Ser* mutation, of whom 32 required beta-blockers and flecainide (age at diagnosis 18 (1-55) years old, start of flecainide at 32 (15-66) years old, 16 (50%) male patients). 47% had an implantable cardioverter defibrillator (ICD). Flecainide was indicated for ventricular arrhythmia (97%) during the exercise test despite beta-blocker therapy and median treatment duration with flecainide was of 7.3 years. All patients were on propranolol (median dose 65 mg/day). Flecainide (median dose 100 mg/day) was well tolerated, with no syncope or stress-induced dizziness. Before flecainide, 5 patients (16%) had ventricular arrhythmic events in the ICD, with 2 requiring appropriate shocks. After flecainide, no events occurred. Both qualitative (2.07±0.77 vs. 1.22±1.08, p<0.001) and quantitative (69.78±83.17 vs. 15.29±5.53, p<0.001) arrhythmic scores improved significantly after adding flecainide. Additionally, there was a significant increase in METs (Z=-2.564, p=0.010) and a reduction in the maximum heart rate (Z=-2.870, p=0.004) and the percentage of the age-predicted maximum heart rate (Z=-3.403, p=0.001) during the exercise test with the combined therapy.

**Conclusion:** Flecainide and beta-blocker therapy in CPVT patients resulted in significant improvements in exercise capacity and a reduction in arrhythmic burden in the long-term follow-up.

## 1. Introduction

Catecholaminergic polymorphic ventricular tachycardia (CPVT) is a rare but life-threatening genetic channelopathy, primarily affecting young individuals without structural heart disease or ischemia and with a normal baseline electrocardiogram. The tachyarrhythmias are usually precipitated by the adrenergic stimulation of physical or emotional stress, leading to the appearance of syncope or sudden death^1^, reason why the diagnosis must be carried out through exercise testing that induces polymorphic ventricular tachycardia, especially bidirectional, and reproduces the patient’s symptoms. Also, detailed family history may reveal relatives with similar events or unexplained sudden death. Although its prevalence is estimated to be low, approximately 1:10,000 in Europe, the disease is highly lethal^2^. In fact, sudden cardiac death is the first clinical manifestation in up to 30% of untreated individuals under 40 years of age^3^.

Six genes are linked to CPVT, with ∼60% of cases caused by heterozygous gain-of-function mutations in the ryanodine receptor-2 (*RyR2*) ^4,5^. These mutations are associated with cardiomyocyte impaired intracellular Ca2+ handling, leading to premature ventricular complexes or bidirectional ventricular tachycardia, seen on ECG as alternating QRS complexes. Genetic testing is essential for diagnosis and risk assessment.

Standard treatment focuses on the use of nonselective beta-blockers, which remain the first-line therapy. However, for patients with persistent symptoms or arrhythmia despite beta-blocker therapy, flecainide and cardiac sympathetic denervation are recommended as second-line interventions^6^. Flecainide, a class IC antiarrhythmic, has demonstrated efficacy in suppressing the ventricular arrhythmia burden in CPVT patients and therefore should be considered in addition to beta-blockers when control of arrhythmia is incomplete. Moreover, in selected patients who show intolerance to beta-blocker therapy, pharmacological therapy with flecainide alone is an option^7^. For those with undiagnosed CPVT who present with sudden cardiac death, an implantable cardioverter-defibrillator (ICD) has not been associated with improved survival. Instead, the ICD has been associated with both a high rate of appropriate/inappropriate ICD shocks along with other device-related complications. Therefore, strict adherence to guideline-directed therapy without an ICD may provide adequate protection^8^.

The objective of this study was to assess the clinical characteristics, safety, and efficacy of flecainide in genotype-positive CPVT patients receiving beta-blockers plus flecainide treatment in the long-term outcome through serial exercise testing.

## 2. Methods

### 2.1. Population

Genotype-positive CPVT patients across the Canary Islands made up our patient population differentiating between those patients who were responders or not to beta-blocker treatment and therefore required the combination with flecainide. Genetic testing was performed in all patients to determine the type of mutation associated with CPVT. Every patient underwent an electrocardiogram, echocardiogram and stress test to rule out the existence of structural and ischemic heart disease. Age, gender, the existence of cardiovascular risk factors (high blood pressure, diabetes mellitus, dyslipidemia and smoking), ICD carriers, the type and dose of beta-blockers, and the dose of flecainide required were also determined. The follow-up period spanned from December 2007 to October 2024. Inclusion criteria were genetically confirmed CPVT, age over 14 at the time of inclusion, being a non-responder to beta-blocker therapy requiring flecainide addition and expressing a willingness to participate by signing the informed consent form. Conversely, patients were excluded if they were under 14 years of age, had significant comorbidities leading to a life expectancy of less than one year, exhibited intolerance to flecainide treatment, or declined to participate in the study. Ethical approval was granted by our Research Ethics Committee.

### 2.2. Clinical Protocol

At the time of diagnosis, all patients underwent an electrocardiogram, echocardiogram, exercise test, genetic testing, and clinical follow-up. All mutation carriers were offered a follow-up and treatment plan, which included a periodic exercise stress test every 3 to 6 months. Additional tests were performed as needed, based on the treating clinician’s assessment. Treatment began with a non-cardioselective beta-blocker, preferably propranolol, at an escalating dose. The treatment goal was to keep the heart rate below 80% of the theoretical maximum for the patient’s age and achieve complete arrhythmia suppression. For carriers in whom arrhythmia suppression was not achieved with the maximum tolerated beta-blocker dose, oral flecainide was added until the therapeutic target was reached. The analysis focused on patients who received flecainide without altering their baseline beta-blocker dosage. Flecainide was initiated with two daily doses, gradually increasing to the maximum tolerated dose or a maximum of 200 mg/day.

### 2.3. Symptom and Arrhythmic Event Definition

Symptoms were defined as syncope (non-vasovagal) or dizziness triggered by physical or emotional stress. Arrhythmic events included appropriate ICD shocks, syncope, or sudden cardiac death.

### 2.4. Definition and Quantification of Ventricular Arrhythmias

Ventricular arrhythmias were categorized as no arrhythmias, isolated ventricular extrasystole, bigeminy, couplets, non-sustained ventricular tachycardia and sustained ventricular tachycardia. Exercise test-induced ventricular arrhythmias was quantified using two scoring systems as seen in Table 1. The Ventricular Arrhythmias Qualitative Score^9^ categorizes arrhythmias into general types, assigning fixed points for each type (e.g., no arrhythmias, couplets, or sustained ventricular tachycardia). It’s more about the presence and type of arrhythmia. On the other hand, the Ventricular Arrhythmias Quantitative Score^10^ is more detailed and assigns points based on the number and frequency of arrhythmic events. For example, it gives different points for isolated ectopic beats, bigeminy, and multiple types of ventricular tachycardia, with higher scores for more frequent or severe events. The quantitative score is thus based on the amount or severity of the arrhythmia, while the qualitative score is based on their general classification.

**Table 1.** Comparison of Qualitative and Quantitative Scores for Each Criterion in the Quantification of Ventricular Arrhythmias.

| Ventricular Arrhythmias | Qualitative Score <sup>(9)</sup> | Ventricular Arrhythmias | Quantitative Score <sup>(10)</sup> |
| --- | --- | --- | --- |
| No arrhythmias or less than 10 EV | 1 | No arrhythmias | 0 |
| More than 10 EV or bigeminy | 2 | Isolated EV | 1 |
| Couplets | 3 | Bigeminy | 15 each |
| Ventricular tachycardia | 4 | Couplets | 30 each |
| Non-sustained ventricular tachycardia |  | Non-sustained ventricular tachycardia | 60 each |
| Sustained ventricular tachycardia |  | Sustained ventricular tachycardia | 120 each |

EV: Ventricular Extrasystole. This table combines both the **Qualitative** and **Quantitative** scoring systems in a single side-by-side format for easy comparison.

### 2.5. Exercise Tests

Exercise tests were conducted using a treadmill with the BRUCE protocol. The exercise test performed before the initiation of flecainide was compared with the exercise test performed during the follow-up at maximum doses of flecainide, without any changes to the beta-blocker treatment. All patients underwent a qualitative and quantitative analysis of exercise test which included all the ventricular arrhythmias detected. To calculate the percentage of age-predicted maximum heart rate (APMHR) we used the formula: [APMHR = (maximum heart rate achieved x 100)/ theoretical maximum heart rate], where the theoretical maximum heart rate was calculated as 220 – age in years.

### 2.6. Statistical Analysis

Data were examined to assess normality using the Kolmogorov-Smirnov test and variables were expressed as mean ± standard deviation or as median and percentiles (5-95) depending on whether they followed a normal distribution or not, respectively. The Wilcoxon signed-rank test was used to compare the differences between paired observations. The Z statistic was calculated to assess the magnitude of the difference, and its significance was evaluated by comparing the Z value to the critical values from the standard normal distribution. A p-value of less than 0.05 was considered statistically significant. The statistical analysis was performed using IBM SPSS Statistics (version 25, IBM Corp., Armonk, NY, USA).

## 3. Results

Of the 235 patients in our series with CPVT, all of whom tested positive for a pathogenic mutation in the gene encoding ryanodine (*RyR2 p.Gly357Ser*), 40 died of sudden death, 12 of non-cardiovascular death, 77 were lost to follow-up, and 1 refused treatment. Of the remaining 105 patients, 73 were receiving beta-blockers and 32 were receiving beta-blockers plus flecainide. Of the latter, 15 were ICD carriers and 17 were not. Figure 1 shows the flow chart of the patients in our series.

**Figure 1.**
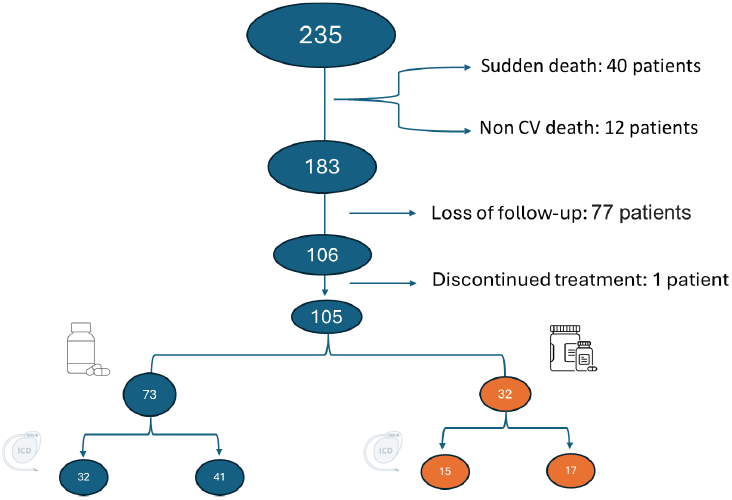
Flow chart of the patients in our series. On the left of the figure patients under beta-blocker treatment and on the right (in orange color) patients under beta-blocker and flecainide therapy. ICD: Patients with implantable cardioverter defibrillator (ICD).

Therefore, our study cohort comprised 32 patients with CPVT (age at diagnosis 18 (1-55) years old, start of flecainide at 32 (15-66) years old, 16 (50%) male patients) all of whom were genotype-positive for the *RyR2 p.Gly357Ser* mutation. None of the patients had a history of arterial hypertension, diabetes, dyslipidemia, or smoking, and all were genetically linked to positive relatives with CPVT. Family history revealed that 41% had a history of sudden death or syncope. Most patients (66%) were asymptomatic at the time of inclusion, while 25% had an arrhythmic syncope and 9% experienced recovered sudden cardiac death. Treatment included beta-blocker therapy with propranolol (median dose 65 mg/d) and flecainide (median dose 100 mg/d). Flecainide was prescribed primarily for ventricular arrhythmias despite beta-blocker treatment (97%). The median duration of flecainide treatment was 7.3 years (range 1-13.6). Almost half of the patients (47%) had an ICD, but none underwent left-cardiac sympathetic denervation. The median number of exercise tests performed during flecainide treatment was of 16, ranging from 2 to 35. No patient had syncope, dizziness triggered by physical or emotional stress during the follow up and all of them had a good tolerance to flecainide therapy. In the pre-flecainide stage, 5 patients (16%) experienced ventricular arrhythmic events recorded by the ICD: 2 patients received appropriate shocks due to polymorphic ventricular tachycardia at over 220 beats per minute, and 3 patients had sustained ventricular tachycardia. However, after initiating treatment with flecainide, no ventricular arrhythmic events were recorded. Table 2 summarizes clinical data in CPVT patients under beta-blocker and flecainide treatment.

**Table 2.**
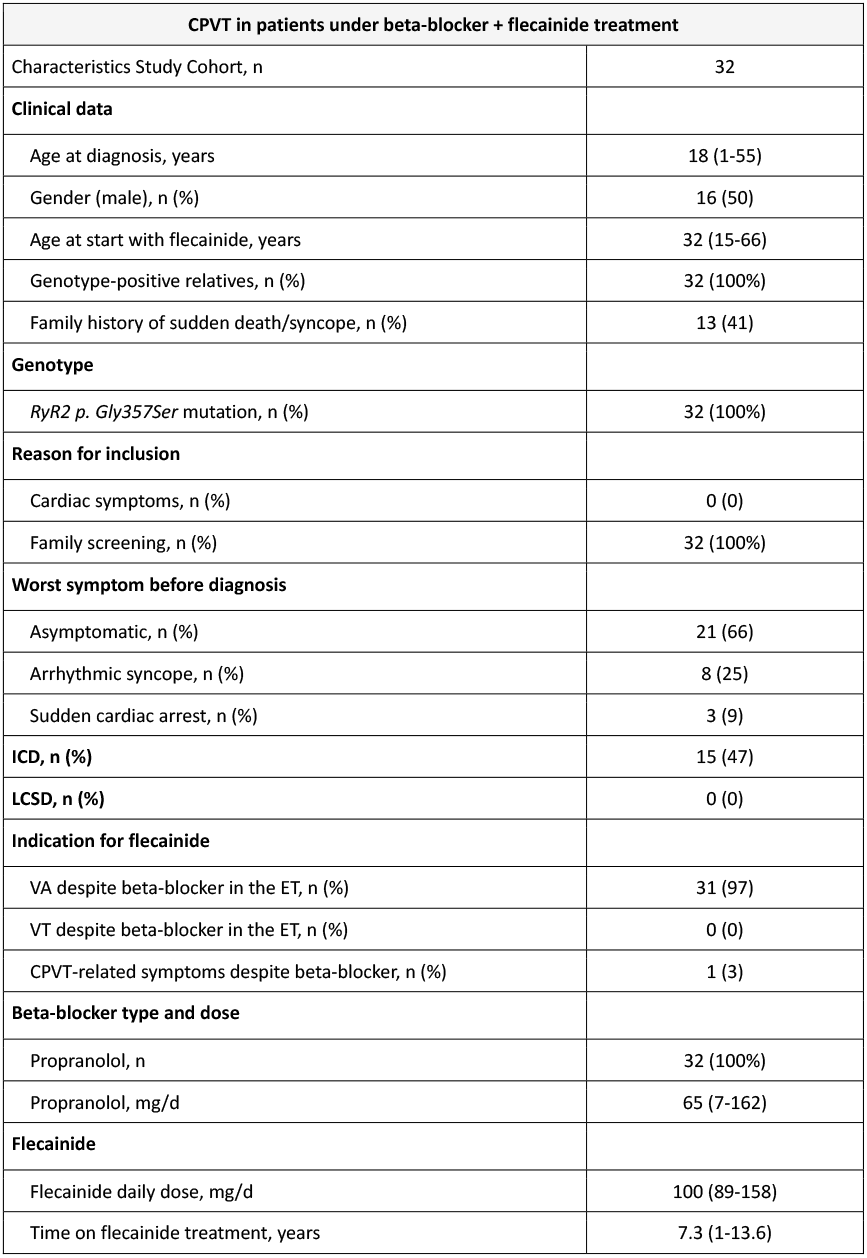
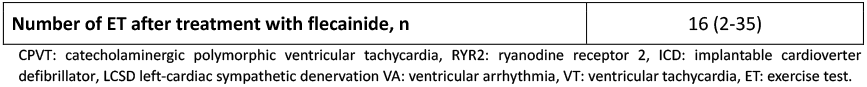
Clinical data of patients with catecholaminergic polymorphic ventricular tachycardia under β-Blocker and Flecainide therapy.

The Wilcoxon test (Table 3) was used to assess differences in treadmill ergometry parameters before and after treatment with flecainide. The results indicate that baseline heart rate did not show significant changes after treatment (Z=-0.216, p=0.829), suggesting that flecainide did not affect resting heart rate. In contrast, significant reductions were observed in the maximum heart rate achieved (Z=-2.870, p=0.004) and the percentage of the APMHR during the exercise test (Z=-3.403, p=0.001) indicating that the treatment with flecainide helped to control the heart rate response during exertion. Additionally, METs increased significantly after treatment (Z=-2.564, p=0.010), reflecting an improvement in patients’ functional capacity, likely associated with greater cardiovascular efficiency. On the other hand, both the qualitative score (Z=-4.152, p<0.001) and the quantitative score (Z=-4.002, p<0.001) decreased significantly, indicating an improvement following flecainide treatment. Figure 2 shows a graphical representation of the data obtained in Table 2 comparing treadmill test media data before and after starting flecainide therapy.

**Table 3.** Exercise test data before and after initiation of flecainide therapy.

| **Exercise Test** | **Before Flecainide** | **After Flecainide** | **Z** | **p** |
| --- | --- | --- | --- | --- |
| Resting heart rate, bpm | 62.38 ± 12.15 | 62.12 ± 9.50 | -0.216 | 0.829 |
| HRmax, bpm | 138.57 ± 22.72 | 128.42 ± 13.00 | -2.870 | 0.004 |
| APMHR, % | 73.54 ± 12.49 | 64.97 ± 13.17 | -3.403 | 0.001 |
| METs | 12.31 ± 3.83 | 14.67 ± 7.48 | -2.564 | 0.010 |
| Qualitative score | 2.07 ± 0.77 | 1.22 ± 1.08 | -4.152 | <0.001 |
| Quantitative score | 69.78 ± 83.17 | 15.29 ± 5.53 | -4.002 | <0.001 |

bpm: beats per minute, HRmax: maximum heart rate achieved, APMHR: age-predicted maximum heart rate, METs: metabolic equivalents. Qualitative<sup>9</sup> and Quantitative<sup>10</sup> Scores for the quantification of ventricular arrhythmias. "Before Flecainide" refers to the exercise test performed on beta-blocker therapy before starting treatment with flecainide while "After Flecainide" refers to all exercise tests performed under beta-blocker and flecainide treatment.

**Figure 2.**
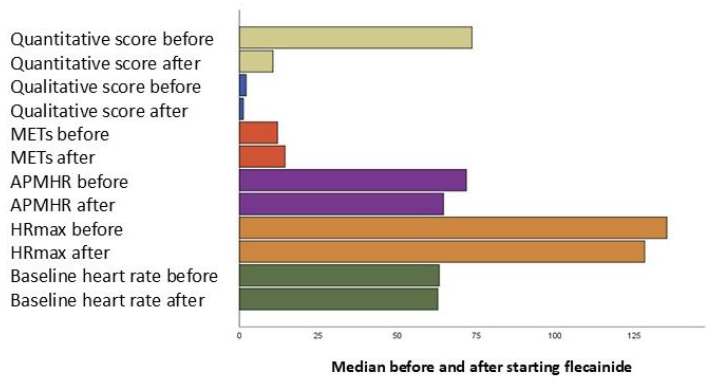
Graphical representation of exercise test data before and after flecainide treatment. Qualitative^9^ and Quantitative^10^ Scores for the quantification of ventricular arrhythmias. METs: metabolic equivalents. APMHR: age-predicted maximum heart rate (in percentage), HRmax: maximum heart rate achieved (in beats per minute). Baseline heart rate is measured in beats per minute. “Before” refers to the exercise test performed on beta-blocker therapy before starting treatment with flecainide while “after” refers to all exercise tests performed under beta-blocker and flecainide treatment.

## 4. Discussion

CPVT is a potentially life-threatening condition that significantly impacts the cardiovascular health of affected individuals, particularly in the presence of exercise-induced arrhythmia. Currently, the two main genetic variants associated with CPVT involve the ryanodine receptor 2 (*RYR2*), as occurred in our series, and calsequestrin 2 (*CASQ2*). While *RYR2* variants cause the most common phenotype with autosomal dominant inheritance, which would explain why 100% of our patients had affected relatives, *CASQ2* variants causing CPVT are autosomal recessive or dominant^11,12^.

Early onset ventricular arrhythmias and exercise-induced ventricular tachycardia at baseline exercise stress test have been associated with life-threatening arrhythmic events at follow-up among patients with CPVT, allowing to identify a sub-set of patients at higher risk at diagnosis^13^. The importance of performing serial testing, as we did in our series, lies in the fact that patients, due to factors such as age or hormonal changes, may have an arrhythmic tendency that can only be detected through repeated stress tests. This approach allows for more tailored and personalized medicine, as it considers the specific condition of the patient at different stages of their life, providing a dynamic assessment that adapts to the changes in their health over time. Serial testing helps identify and manage potential arrhythmic risks early, along with the adjustment of the flecainide dose.

The standard approach to managing such patients often involves beta-blockers, which are effective in reducing arrhythmic events and improving patient outcomes. However, there is a subset of patients in whom beta-blockers alone fail to provide sufficient control, necessitating the addition of antiarrhythmic agents like flecainide. Previous studies have evaluated the efficacy and safety of flecainide in addition to conventional drug therapy in patients with CPVT to reduce the arrhythmogenic burden^14^. Kannankeril et al.^15^, for example, in a randomized clinical trial evidenced that flecainide plus beta-blocker significantly reduced ventricular ectopy during exercise compared with placebo plus beta-blocker and beta-blocker alone which is line with our results. However, unlike our study, the number of patients was small; the follow-up period was short, and the number of ergometries was limited. In this context, our study supports the safety and beneficial role of adding flecainide to beta-blocker therapy in the long term follow up of CPVT patients, demonstrating significant improvements in the heart rate, the arrhythmic threshold and the functional capacity during the follow-up ergometries. Additionally, it is important to note that counting premature ventricular contractions on Holter monitors is insufficient for diagnosing CPVT due to the adrenergic nature of the disease^16^. For this reason, exercise testing, widely used in our follow-up, remains the gold standard for monitoring.

Similarly, Bergeman et al.^17^ observed that adding flecainide to beta-blocker in patients with CPVT was associated with a lower incidence of arrhythmic events in the overall cohort, in symptomatic patients, and particularly in patients with breakthrough arrhythmic events. However, this multicenter study, obtained from two international registries, did not carry out a serial ergometric assessment over time as we did in our patients. In fact, in our study, flecainide added to the standard β-blocker therapy, not only did it prevent the occurrence of arrhythmic events, but it also significantly reduced the arrhythmic burden, both quantitatively and qualitatively, detected during the ergometric tests. Flecainide lowered the percentage of the heart rate threshold which may help in managing ventricular arrhythmia, by preventing the heart from reaching high rates that could trigger more severe episodes like sustained ventricular tachycardia or sudden cardiac arrest. In fact, in athletes with previously symptomatic CPVT for whom return to play is being considered, combination therapy with beta blocker and flecainide, and consideration of triple therapy with LCSD, is recommended before return to play, with a goal of optimizing therapy to normalize the exercise stress test (Class of recommendation 1 with a level of evidence with limited data). Moreover, in such patients bigeminal premature ventricular contractions may be acceptable, but couplets or more extensive non-sustained ventricular tachycardia require continued treatment intensification^18^. The findings of our study support the notion that flecainide, when combined with β-blockers, can provide better cardiovascular efficiency in patients with CPVT, including athletes, providing great long-term safety and effectiveness. Hence, the addition of flecainide raises an important clinical question regarding whether the need for ICD replacement or not. In our cohort, 47% of patients had an ICD implanted prior to initiating flecainide therapy. Although we have not removed any of the previously placed ICDs, we can say that after starting treatment with flecainide it has not been necessary to implant any new ones nor have arrhythmic events been observed in ICD carriers.

Additionally, the use of flecainide without beta-blockers raises another possibility: whether flecainide alone could be sufficient in managing CPVT, especially in patients who do not tolerate or respond to beta-blockers. Although no patients in our study were treated exclusively with flecainide, it would be valuable to explore this approach further. In fact, although nonselective beta-blockers remain the cornerstone treatment for CPVT, 10% of patients with CPVT may require a beta-blocker-free treatment strategy^19^.

Regarding the limitations of our study, one of them is the relatively small sample size, consisting of 32 patients. However, the cohort provides valuable clinical insights into the clinical presentation and treatment response in these patients as it is the largest published cohort to date with long-term follow-up and serial ergometric assessments. Another limitation is the use of a single type of beta-blocker, propranolol, across all patients. Although non-selective beta-blockers are commonly prescribed for CPVT and have been shown to be effective in controlling arrhythmic events, there are other beta-blockers available that may offer different pharmacokinetic properties or varying levels of efficacy^21,22^. Nonetheless, no significant differences between propranolol and nadolol have been seen in relation to life-threatening arrhythmic events^23^. Additionally, our study cohort consisted solely of individuals carrying the *RYR2 p.Gly357Ser* mutation, which displays an increased propensity for spontaneous Ca^2+^ release under conditions of store Ca^2+^ overload in addition to reducing *RyR2* protein expression^10,24^. While this mutation shares similarities with other *RYR2* mutations that also affect calcium handling and predispose patients to arrhythmia under stress, we believe these data are applicable to the rest of the patients with CPVT.

In conclusion, our findings support the addition of flecainide to non-selective beta-blockers therapy for improved arrhythmic control and functional capacity in patients with CPVT who do not respond adequately to beta-blockers alone. While the potential reduction of ICD implantation or the use of flecainide without beta-blockers remains an area for further investigation, our study provides promising evidence of the therapeutic benefits of flecainide in managing this complex arrhythmic condition.

## Data Availability

"All data supporting the findings of this study are available from the corresponding author upon reasonable request."

